# Lowering The Acoustic Noise Burden in MRI with Predictive Noise Canceling

**DOI:** 10.1101/2024.04.28.24305337

**Authors:** Paulina Šiurytė, Sebastian Weingärtner

## Abstract

Even though Magnetic Resonance Imaging (MRI) exams are performed up to 16 times per every 100 inhabitants each year, patient comfort and acceptance rates are strongly compromised by exposure to loud acoustic noise. Here we present a system for acoustic noise cancellation using anti-noise derived from predicted scanner sounds. In this approach, termed predictive noise canceling (PNC), the acoustic fingerprint of an MRI system is obtained during a 60 s calibration, and used to predict anti-noise for arbitrary scan procedures. PNC achieves acoustic noise attenuation of up to 13 dB across a wide range of clinical MRI sequences, with spectral noise peak reduction of up to 96.76 % occurring between 0.6 and 1.2 kHz. These results suggest that predicted scanner noise can achieve substantial in-bore noise cancellation with the prospect of providing a cheap and scanner-independent solution for improved patient comfort.

---

THe average person in the USA will have undergone seven medical Magnetic Resonance Imaging (MRI) scans by the end of their life - an increasing figure due to population aging^1^. Despite being known as safe and radiation-free, MRI exposes patients to extreme sound pressure levels (SPL) of up to 130 dB for prolonged time periods^2,3^. This far exceeds public health recommendations^4^ and puts patients at risk of temporary or permanent shift of the hearing threshold^5–8^. Patient discomfort and anxiety, caused by the noise burden, further contribute to poor patient acceptance ratings, well below those of more damaging imaging modalities such as computed tomography^9^. Thus, novel solutions for reducing acoustic noise levels in clinical scanning sites worldwide are essential to ensure both patient comfort and safety.

Here, we evaluate a novel approach to active noise canceling (ANC) in MRI that allows for versatile noise reduction compatible with any MRI system and scan procedure. In the proposed approach, termed predictive noise canceling (PNC), anti-noise is generated prior to the scan procedure, based on the gradient input and a pre-calibrated acoustic noise correlation (see the animated abstract in Supplementary Video S1).. We demonstrate live acoustic noise reduction inside a clinical 3 T MRI scanner using a pneumatic headphone-imitating setup. The effectiveness of PNC is shown for a range of MRI sequences representative of a clinical scan portfolio. The impact of various MRI sequence parameters on the noise reduction capabilities is systematically studied and sources of noise cancelation imperfection are dissected.

## Results

The main source of acoustic noise during MRI scans is the vibration caused by rapidly switching currents in the magnetic field gradient coils (see Figs. 1A-B)^10^. In clinical MRI systems, the acoustic noise lies predominantly in the 0-3 kHz range^11^, where the human ear is highly sensitive^3^ (see Fig. 1C). Potential methods for alleviating the noise burden include passive noise damping with headphones or earplugs^12^, as well as hardware advances such as vacuum-mounted gradient coils^13^ or ultra-fast gradient switching^14^. Passive noise reduction is mandatory in clinical use but leaves the most problematic lower frequencies vulnerable when used alone^4,12,15^ (see Fig. 1C-D). Hardware upgrades are no sustainable solution, as they are often incompatible with existing systems and are too costly for widespread use. ANC has received interest for suppressing low frequency noise. However, the strong magnetic fields in MRI restrict the acoustic equipment that can be used, greatly hindering the effectiveness of ANC^16–18^. For example, pneumatic sound transfer, as most commonly used in commercial MRI intercommunication systems^19,20^, incurs considerable latency, leading to a diminished frequency range for ANC effectiveness. As a result, recently introduced commercial ANC devices^21^, report 30 dB reduction for ANC combined with passive cancellation, which is in line with the effectiveness of passive-only solutions^22^.

**FIGURE 1.**
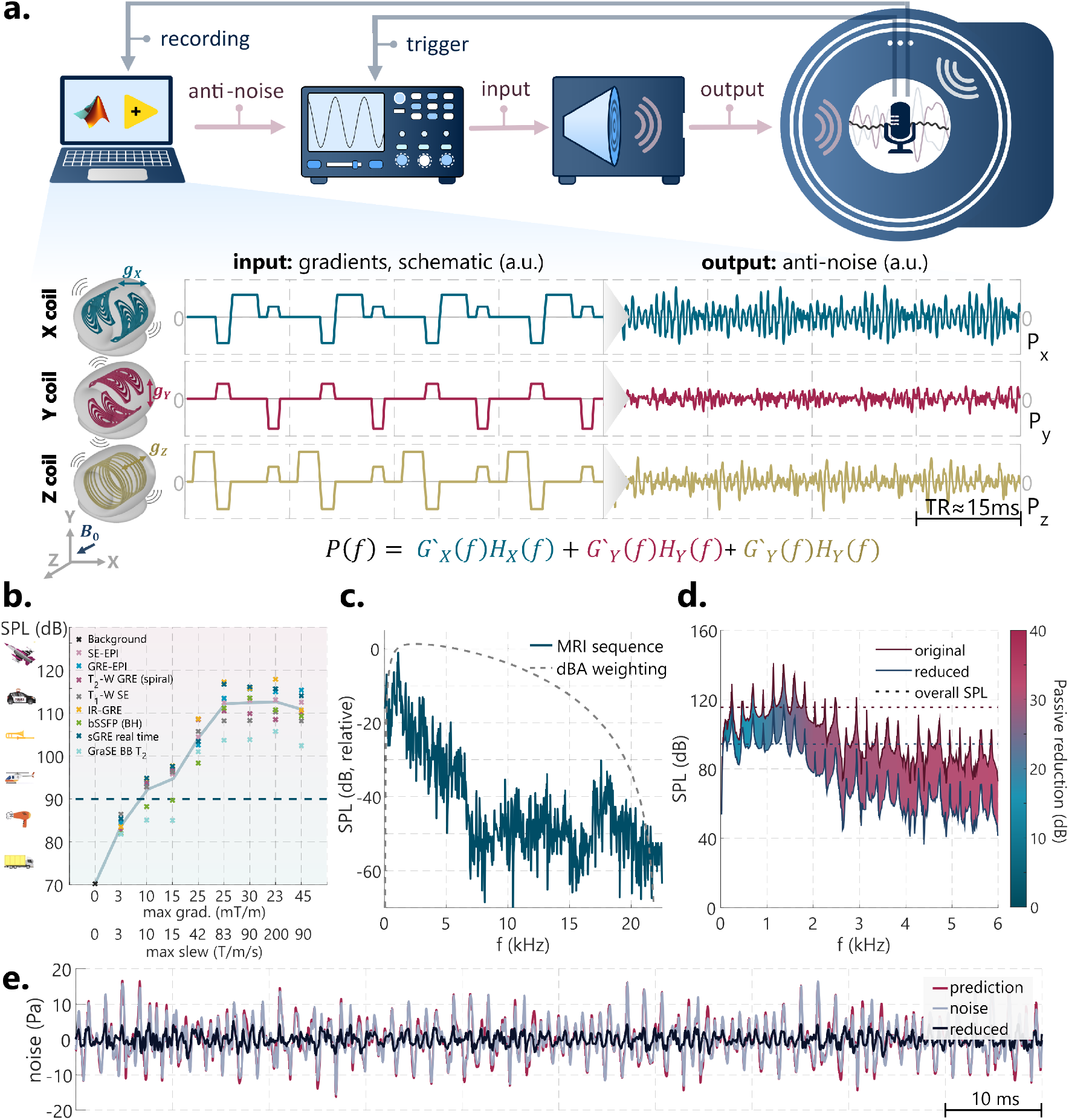
Predictive Noise Canceling (PNC) pipeline and MRI acoustic noise characterization. **a, top**, PNC pipeline, comprising a control PC, an arbitrary function generator (AFG) for synchronized signal production, a sound playback system, and the MRI scanner. Optical microphone recordings are collected and processed, while time synchronization between the signal output of the AFG and the MRI scanner is ensured using a transistor-transistor logic (TTL) trigger. An optical microphone inside the scanner bore picks up the scanner sound for obtaining the acoustic fingerprint of the scanner. **a, bottom**, schematic representation of a representative coil input *g*_*X/Y/Z*_ during a spoiled GRE MRI sequence (TR ≈ 15 ms) and corresponding simulated sequence noise *p*_*X/Y/Z*_. **b**, exacerbating noise burden of selected sequences for increasing gradient coil performance. Blue dashed line at 90 dB indicates the safe limit for prolonged exposure. Gradient system limits are based on historical scanner specifications^23^. **c**, example spectrum of MRI sequence noise (blue) and dBA weighting, approximating human hearing sensitivity (dashed line). **d**, example sequence noise spectrum before (pink line) and after (blue line) the application of passive reduction (earplugs), indicating the weakest reduction at lower frequencies. The dashed lines indicate overall noise reduction.

### MRI-compatible noise canceling setup

To demonstrate latency-invariant noise reduction using prediction-based anti-noise, an experimental setup was constructed. An optical fiber microphone is placed inside the MRI scanner (Ingenia 3T, Philips Healthcare, Amsterdam, The Netherlands). Sound is transmitted to the in-bore position from an amplifier-driven speaker outside the scanner room through a pneumatic hose and funnel. Precise sound waves are generated by an arbitrary function generator (AFG) that powers the amplifier. The AFG is controlled using an external TTL (transistor-transistor logic) trigger from the MRI control system, to synchronize the MRI noise and the antinoise with high temporal precision. A detailed description of the components and specifications of the setup is provided in the Methods section and Fig. 1A.

A linear time-invariant (LTI) model is used to predict the total X/Y/Z gradient coil noise from the derivative of the gradient current inputs. Details evaluating the suitability of the LTI model are provided in the Supplementary Materials and Figure S1. To construct the LTI model, individual transfer functions are derived for each of the three gradient coils, from a 60 s calibration procedure with triangular gradient pulses. This calibration procedure is setup-specific for a given subject positioning and needs to be performed only once for a conventional scanning session.

All noise cancellation experiments are assessed in two frequency ranges: a broader spectrum (0.3-4 kHz), encompassing the majority of MRI sequence noise^3^, and a narrower range (0.5-2 kHz), targeting the most intense sound pressure level (SPL) regions^24^. The noise reduction is described in the wide frequency range if not specified otherwise.

### Feed-forward signal corrections

Feed-forward corrections are applied to the anti-noise prediction for pre-emptive compensation of inaccuracies in the sound playback system. For our setup, three dominant error sources are considered: 1) channel distortion, 2) output latency, and 3) recorder clock mismatch. All corrections are derived from on the 60 s calibration procedure only.

Channel distortion, as a result of the frequency response of the speaker and the sound transmission through the pneumatic hose, causes a mismatch between the intended and actual output (Fig. 2A). Equalization (EQ) of the output was performed in multiple iterations using linear inverse-distortion filters derived from gradient blips in the calibration sequence (Fig. 2B). Without EQ, live noise reduction showed negligible 0.23±0.62 dB noise cancellation across all three gradient coils. Applying EQ with an increasing number of iterations lead to improved live noise reduction, approaching a plateau at 18.62±1.11 dB with three EQ iterations (Fig. 2C). Thus, three-iteration EQ was used for the remainder of the study.

**FIGURE 2.**
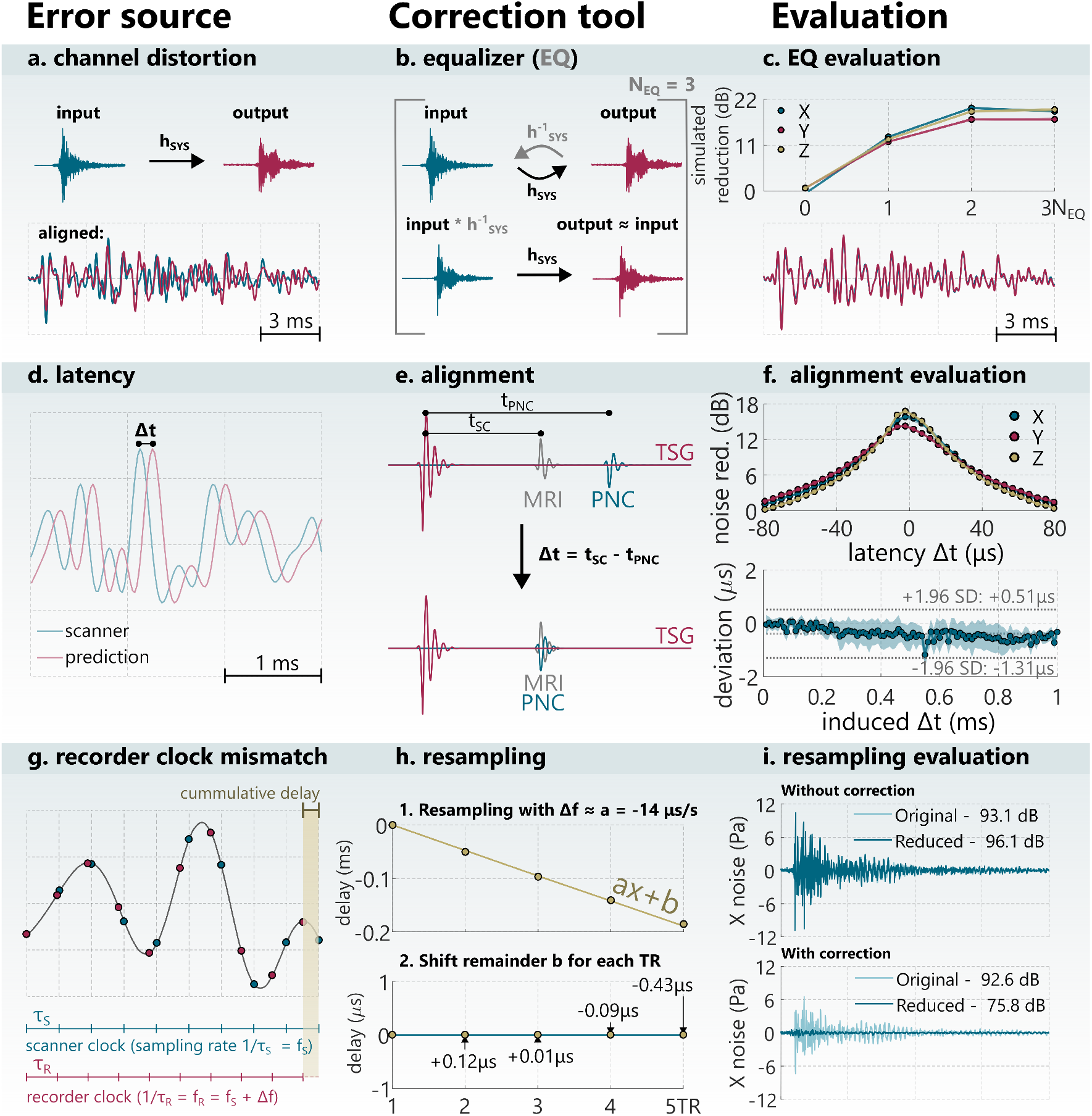
Feed-forward corrections for effective noise cancellation. **a**, channel distortion is described by the transfer function, *h*_*sys*_ (top), and leads to deviation from the desired output (bottom). **b**, equalization (EQ) via inverse-distortion filters, 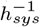. **c**, exemplary noise reduction for different EQ iterations shows convergence at 3 iterations (top), leading to minimal output distortion (bottom).**d**, latency incurred during processing, sound production and transport. **e**, latency Δ*t* is estimated from the relative delay between the TTL synchronized gradient (TSG) sound and MRI gradient blip and the equalized PNC output. **f**, pronounced latency lead to poor noise reduction of triangular gradient pulses (0.14 ms rise time and 20 mT/m amplitude) (top), but retrospectively induced latency shows that it can be accurately estimated and corrected for (bottom).**g**, recorder clock-mismatch is apparent in PC recordings as subsequent TR pulse dephasing (top). This results in approximately linear cumulative latency (bottom). **h**, sampling mismatch is corrected by linear resampling (top), followed by individual sample point shifts applied using an estimation of the remainder latency, *b*_*n*_. **i**, live reduction examples with/without the correction (top/bottom) for a triangular X gradient (0.14 ms rise time and 20 mT/m amplitude) pulse (bottom), show that sampling correction is necessary for effective noise cancellation.

Digital processing delays and sound travel times induce latency in the sound system. Pre-emptive latency correction was performed by measuring the delay between the scanner pulse and equivalent equalized output, following a TTL-trigger synchronized gradient blip (TSG) (Fig. 2E). To evaluate the effect of latency on live noise reduction, delays from -80 to 80 *μ*s were induced in the anti-noise predicted for X/Y/Z gradient pulses. PNC reduction efficiency in decibels was halved for 30.70±4.29 *μ*s latency in triangular gradient blip experiments (Fig. 2F). In a separate experiment, increasing delay was induced between the TSG and a gradient blip, ranging from 10 to 1000 *μ*s in steps of 10 *μ*s. Bland-Altman analysis of the induced and retrospectively measured delays shows less than 1.19 *μ*s deviation. This suggests less than 0.5 dB expected loss in noise reduction efficiency due to residual latency effects.

Finally, the sampling rate of the recorder clock shows deviations from the scanner clock, leading to a cumulative delay (Fig. 2G). Linear resampling was applied globally to all recorded signals, based on the recording drift of repeated TSG pulses (Fig. 2H, top). Additionally, the residual short-term recorder clock variations in the calibration sequence were eliminated by shifting the samples of the individual gradient blip sounds (Fig. 2H, bottom). Live noise reduction for X coil gradient blips without the clock-mismatch correction achieved poor reduction performance (3.08±0.03 dB) due to dephasing of averaged calibration pulses. With correction, 16.87±0.04 dB live reduction was achieved (Fig. 2I).

### Up to 13 dB noise reduction in MRI sequences

A test protocol of ten MRI sequences (Supplementary Table S1), chosen to be representative of modern clinical MRI scan methods, was performed with and without PNC. Experiments were repeated in six scanning sessions, each acquiring one reference without PNC, one prediction-only recording, and five repetitions with PNC for each sequence. PNC achieves consistent sound pressure reduction for all sequences, as illustrated in representative noise recordings in the time and frequency domain in Figure 3A (audio-visual illustration in Supplementary Videos S2-11). Up to 12.65 dB reduction in the wide frequency range and up to 13.55 dB in the narrow range were observed in individual sequence experiment iterations. The frequency spectrum of the sequences shows peak SPL between 600 and 1170 Hz, where noise reduction achieves 55.91-96.76% sound pressure reduction. Across all sequences and all scanning sessions, the mean reduction was 9.21±1.23 dB for the wide frequency range and 9.97±1.48 dB for the narrow range. For individual sequences, this ranged from 7.98±0.58 to 10.01±1.04 dB and from 8.75±1.73 to 11.00±1.39 dB in wide and narrow considered ranges, respectively (Fig. 3B).

**FIGURE 3.**
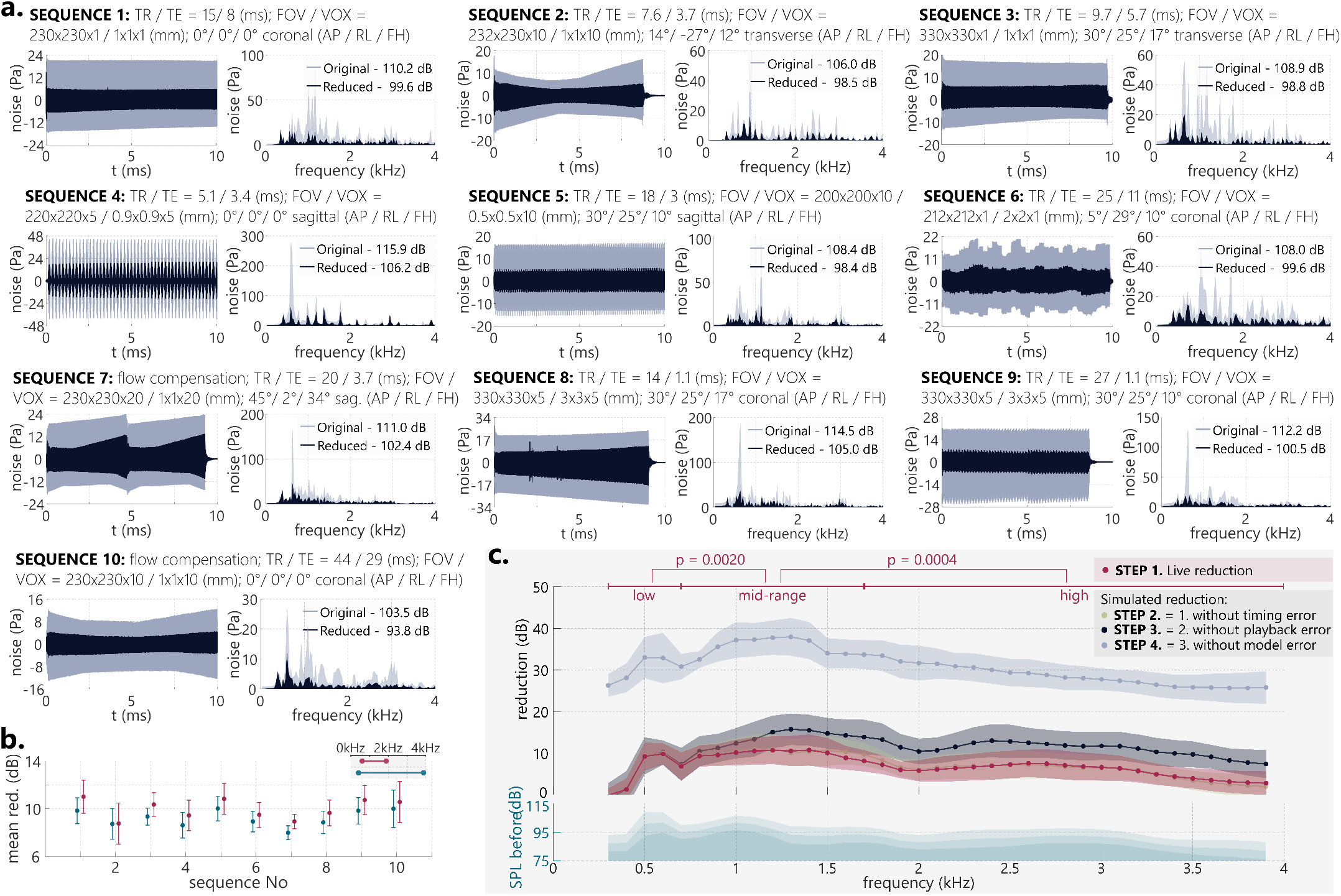
Acoustic noise reduction in representative MRI sequences. **a**, sound pressure levels of the acoustic noise in time (left) and frequency domains (right) with and without PNC application. Sequences are characterized by repetition and echo time (TR/TE), field of view and voxel size (FOV/VOX), orientation, and anterior-posterior/right-left/feed-head angulation (AP/RL/FH). The legends indicate the overall reduction in 0.3-4 kHz range **b**, a representative sequence noise reduction results, evaluated in wide 0.3-4 kHz range (blue) and narrow 0.5-2 kHz range (pink). **c**, dissection of the acoustic noise reduction in the frequency spectrum, showing live noise reduction and simulated reduction, when virtually eliminating error sources related to timing, sound playback, and model limitations. Data points represent 100 Hz bins. PNC shows significantly higher noise reduction in the mid-frequency range compared with low/high frequencies.

To dissect the noise cancellation results, four experimental error sources were isolated step-wise:

**Step 1:** measured live noise reduction;

**Step 2:** simulated reduction without timing error, using retrospective noise/anti-noise alignment;

**Step 3:** simulated reduction as in step 2, but without playback error, using pre-output prediction as anti-noise;

**Step 4:** simulated reduction as in step 3, but without the LTI model error, using pre-recorded sequence noise as prediction.

The most significant reduction in live noise occurs within the mid-frequency range (700-1700 Hz), with statistically significant differences compared to both low frequencies (300-700 Hz, *p* ≤ 0.002) and high frequencies (1700-4000 Hz, *p* ≤ 0.0004). Residual latency minimally affected noise reduction (Step 2), indicating highly effective latency control. Imperfect channel equalization (Step 2, compared to Step 3) reduced the effectiveness of PNC mostly at higher frequencies by 5.27±0.57 dB. Simulated noise reduction under idealized conditions (Step 4) yields up to 40 dB (Fig. 3C), indicating the largest efficiency loss is due to the LTI model imperfections. The same conclusions can be drawn from the noise reduction analysis for isolated gradient sounds, as shown in Fig. S2.

### Live reduction is robust to sequence modifications

Trends in the performance of PNC were tested by varying a range of acoustically relevant sequence parameters: repetition time (TR = 6.45 - 550.40 ms), slice-thickness (0.5 - 10 mm), bandwidth (BW = 271.7 - 617.3 Hz/Px), and slice angulation (angle = 0° - 162°). An exemplary balanced Steady State Free Precession (bSSFP) was used as base sequence (see Supplementary Table S1). For each parameter setting, noise reduction in wide and narrow frequency ranges was obtained from recordings with PNC in five repetitions and one reference measurement without PNC.

Thicker slices, leading to weaker slice selection gradients, show increased noise reduction with up to 11.00±0.12 dB (Fig. 4A, 10 mm). At very thin slices (0.5 mm) (Fig. 4A), noise reduction was compromised with 6.23±0.12 dB compared to 10.58±0.64 across 1-10 mm slices.

**FIGURE 4.**
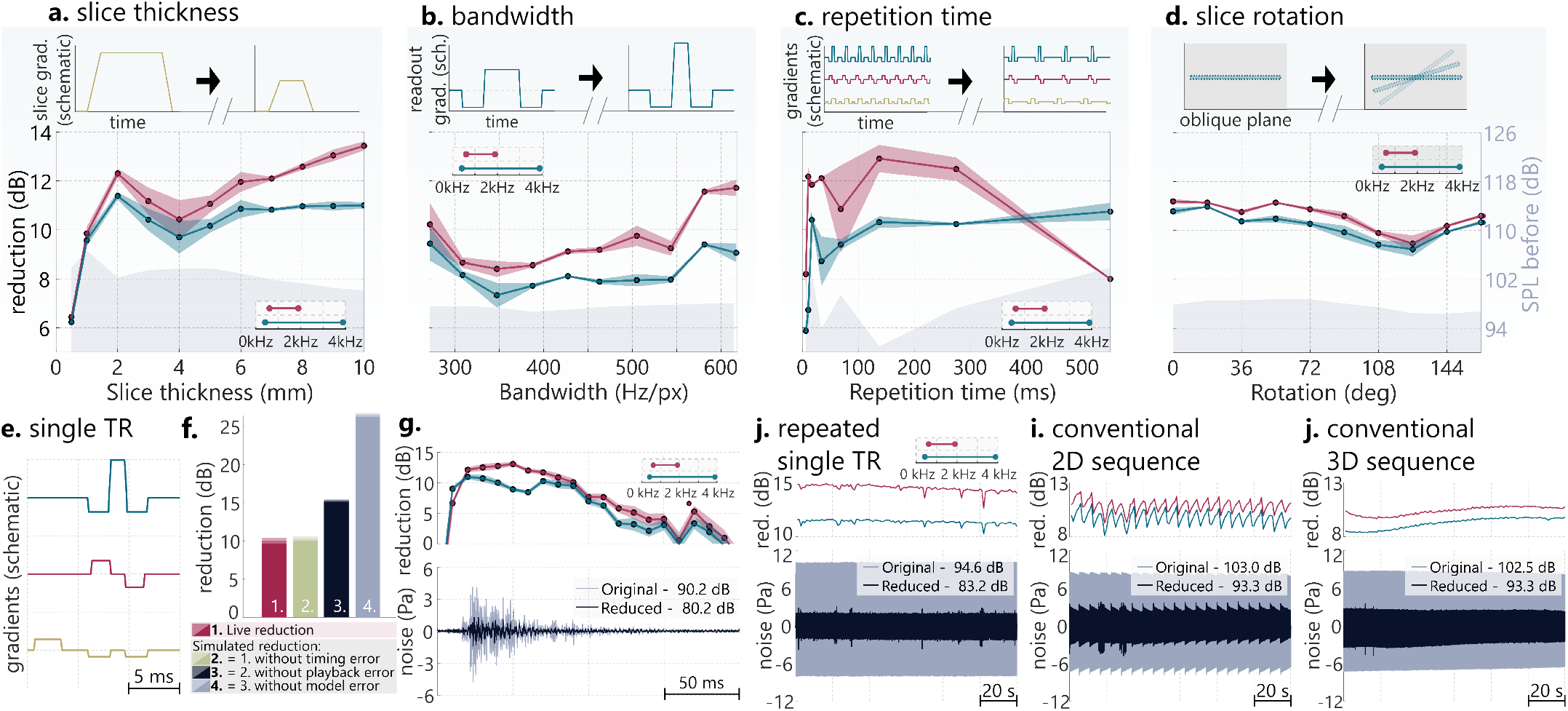
Noise reduction for variations of acoustically relevant MRI sequence parameters. **a-d**, noise reduction in 0.3-4 kHz (blue) and 0.5-2 kHz (pink), with shading indicating the standard deviation. Gray shading depicts the sound pressure level (SPL) without reduction., slice thickness, affecting slice selection gradient; **b**, bandwidth, affecting the readout gradient; **c**, repetition time, affecting gradient spacing over time; **d**, slice rotation, affecting gradient load distribution across physical coils. All gradient waveforms are schematic representations. **e-g**, noise reduction analysis on short time scale, showing **e**, representative schematic single TR gradients of a conventional MRI sequence; **f**, short time scale analysis of a single TR clip, with live noise reduction and simulated reduction when eliminating errors related to timing, sound playback, and modeling; **g. h-j**, noise reduction across long time scales, evaluated across a 1 s sliding window (top) in 0.3-4 and 0.5-2 kHz ranges, and an overlay of original and reduced noise over time (bottom). **h**, identical single TR clip from **e** repeated over 2 minutes, **i**, regular 2D MRI sequence repeated in multiple dynamic repetitions over 2 minutes, **h**, regular single 3D MRI scan with a 2 minute duration. All long acquisitions show temporally stable noise reduction.

The readout gradient was modified by adjusting the acquisition bandwidth (Fig. 4B). A slight drop in noise reduction was observed for mid-range bandwidths (300 - 450 Hz/Px) with 7.82±0.43 dB, while the best noise reduction was achieved at high bandwidths (> 550 Hz)/Px with 9.23±0.31 dB, where the readout gradient causes a higher overall SPL of the sequence.

Sequence TR was modified to alter the spacing of the gradient pulses, and, therefore, the primary noise frequency. Increasing TR lead to increased noise reduction with 10.75±0.36 dB at the longest considered TR of 550.4 ms (see Fig. 4C). Compromised noise reduction was observed at ultra-short TRs with 5.87±0.01 dB.

Finally, rotating the imaging slice around a double-oblique axis varies the gradient input load across the three physical gradient coils. The observed effect on live noise reduction showed minor variations of up to 1.74±0.29 dB across the orientations. The overall SPL of the sequence without noise reduction was lowest for 108°-144° indicating a differential acoustic response of the three gradient coils (see Fig. 4D).

### Noise reduction remains stable across time

Next, PNC performance stability across time was analyzed with three 2-minute acquisitions: 1) A single-TR clip (12 ms) repeated with 250 ms spacing in 480 repetitions; 2) The corresponding clinical 2D bSSFP sequence (TR = 12 ms) with 21 repetitions of each phase-encoding step; 3) an equivalent length 3D bSSFP sequence (TR = 12 ms) (details in Supplementary Table S1). The single-TR clip was selected as the phase encoding step with maximum gradient strength (see schematic in Fig. 4E).

For an isolated single-TR clip, the mean reduction across the four steps was 10.07±0.37, 10.33±0.31, 15.28±0.15 and 26.48±0.22 dB at steps 1 to 4, respectively, in the wide frequency range (see Fig. 4F). Visualizing the noise reduction within the TR with a 10 ms sliding time window analysis shows that the reduction is most effective at the time point of the peak gradient noise response. Up to 10.96±0.60 and 13.07±0.31 dB reduction was measured in the wide and narrow frequency ranges with a 10 ms sliding window, respectively (Fig. 4G). For the 2-minute experiments, good temporal stability of noise reduction was observed over a 1 s sliding window (Fig. 4H-J). Mean sequence reduction in the first 10 seconds was averaging 11.42±0.18 / 14.69±0.15 dB, 10.00±0.57 / 11.39±0.51 dB and 8.43±0.04 / 10.06±0.18 dB in the wide/narrow ranges for the fixed-phase, 2D and 3D sequences, respectively. Comparatively, in the last 10 seconds, the average reduction was 11.22±0.10 / 14.19±0.09 dB, 9.41±0.54 / 10.76±0.44 dB and 9.70±0.07 / 10.67±0.04 dB. The overall comparable noise performance indicates high temporal stability, with most effective attenuation in SPL-intensive frequency regions.

## Discussion

In this study, predictive generation of anti-noise, based on the gradient coil inputs, was used in a model setup, demonstrating versatile attenuation of MRI sequence sounds of up to 13 dB. Across a wide range of sequences, up to 96.76% sound pressure peak reduction was achieved in live noise reduction experiments. The method showed adaptability to changes in the sequence input and length, requiring only a single calibration for a set patient position per scan session.

The noise prediction in PNC is based on the acoustic modeling of scanner sounds. LTI models have previously been shown to be good candidates for approximation of the gradient coil noise in MRI, in the context of SPL prediction for sequence design^25–29^. Our results demonstrate that an LTI model provides a viable starting point for the predictive anti-noise application, where time domain sound wave match is critical. At the same time, the dissection of the noise reduction performance shows that model limitations are the primary factor of incomplete noise reduction. While good linearity of individual gradient pulses has been shown, gradient superposition and time-invariance violations, such as those induced by gradient heating, may limit the noise reduction capabilities with LTI model. Thus, the use of non-linear, data-driven, or thermal^30^ modeling may provide an avenue to more accurately predict the gradient noise in scan setup-specific ways and further improve the noise reduction capabilities of PNC.

In our experiments, a comprehensive 60 s calibration procedure was designed to enable thorough corrections of error sources in a proof-of-principle setup. Triangular gradient pulses have been used to yield maximum acoustic response across a broad frequency range. For practical use, shorter calibration sequences can be evaluated, for example, using shorter pulse separation, employing a sequence-specific pre-scan, or re-purposing already present pre-scans for noise canceling calibration. Repeated calibration may also be useful in the presence of motion. Due to differences in the wavefield, displacement of the head in the centimeter range may lead to significant reduction, which can be alleviated with re-calibration.

To demonstrate the effectiveness of PNC in-situ, a pneumatic single-channel headphone model was constructed. This resulted in additional unwanted errors in the signal output. The four-step reduction cascade analysis revealed that after including LTI model, channel distortion was the biggest experimental contributor to reduced noise canceling. While careful equalizer (EQ) calibration achieved up to 18.62±1.11 dB simulated reduction, deep nulls in sound transmission may be present, for instance, as a result of the wave transduction through the hose. Steady improvements in high-fidelity hardware have enabled MRI-compatible acoustic devices with excellent acoustic properties. For example, piezo-electric speakers^31^, electro-dynamic devices^32^ or micro-electro-mechanical system (MEMS)^22^ have been proposed for clear sound production in two-channel headphones. The integration of PNC with these hardware developments bears great promise for improved reduction capabilities and warrants investigation in future studies.

In clinical MRI, the predominant cylindrical scanner shape acts as a waveguide and causes resonances at certain frequencies^11^. In addition, room dimensions result in acoustic echoes, presenting secondary sound waves. PNC can offer a location-specific solution adaptive to the overall room acoustics. While the majority of acoustic noise is perceived through air sound pressure waves, residual noise burden is perceived from the vibrations of bone and tissue. This transmission path is ≈40 dB less efficient^11^, but can be enhanced at low frequencies with the use of earplugs via the occlusion effect^33^. Headphone-based application of anti-noise may have limited effect on reducing the noise burden experienced from bone conduction. However, a subject-specific adaptation of the anti-noise has previously been reported to lower the psycho-acoustic noise burden from bone conduction^34^. Furthermore, in PNC, the creation of larger silent zones may be attainable if speaker arrays are used, instead of headphones. Creating silent zones around the skull or relevant patient areas in other applications, such as neonatal imaging, may therefore be a promising pathway to tackle residual bone conduction in a PNC setting.

PNC showed up to 13 dB reduction In the 0.3-4 kHz range in our experiments. While hardware-related approaches to noise reduction boast up to 30 dB^13,14^ and sequence-tailoring may reach 20-40 dBA reduction^3^, these approaches require either costly upgrades or trade-offs in acquisition quality. PNC could offer an affordable add-on solution to substantially reduce the acoustic noise in MRI without altering the imaging process. While the pneumatic system with PNC in this work achieved on average lower reduction than reported ANC values of 10-30 dB^24^, it showed peak performance in the SPL-intense mid-frequencies. ANC approach attenuation is most suitable for low frequencies below 700 Hz^3^, offering less overlap with the SPL-intense regions of MRI sequence noise. Combining PNC with ANC and passive reduction methods could provide a particularly balanced reduction spectrum, with effective attenuation throughout low, mid-range and high frequencies.

PNC is intrinsically compatible with a wide range of scan systems. Gradient noise is known to scale up with the main magnetic field due to increased Lorentz forces^35,36^ despite Lorentz damping^11^, leading to concerns about SPL in emerging ultra-high field applications. Gradient amplifier improvements enable increasingly higher slew rate and amplitude capabilities. In the absence of better gradient assembly damping, this leads to louder acoustic output (Fig. 1B). The resulting noise burden strongly contributes to severe anxiety and claustrophobic reactions, which occur in 5-10 % of patients^37^. Thus, implementation of acoustic noise reduction methods is indispensable and timely, and together with other comfort-oriented solutions may reduce claustrophobic incidents by up to threefold^38^. In this light, predictive noise canceling bears great promise as a flexible and cost-effective solution for lowering the acoustic noise burden in existing and new MRI sites.

## Methods

### Experimental setup details

The headphone model consisted of a single-channel optical fiber microphone (Phonoptics, France), placed inside the bore next to a pneumatic rubber hose (3.5 cm inner diameter, 7 m long) with a widening plastic funnel end (20 cm diameter). To maintain microphone position stability across scanning sessions, a half-cone frame-shaped holder was printed with polylactic acid (PLA) and mounted on the funnel. The microphone was manually positioned at the isocenter of the MRI scanner. In the control room, the hose was connected to a widening funnel inside a custom-built speaker box with a fitted 20 cm diameter woofer. The speaker box was connected to a signal amplifier (Technics, Japan). The microphone signal was recorded with a PC system (Windows 11, 11th Gen Intel(R) Core(TM) i7-1165G7 CPU) at 44.1 kHz sampling rate. Anti-noise was played through an AFG (AFG31002, Tektronix, US), providing high temporal resolution (250 MHz sampling frequency). For time synchronization with the MRI, an external TTL signal was passed and received by the AFG to trigger the output using the Functional Brain Imaging Box (Philips Healthcare, Amsterdam, The Netherlands).

All experiments were performed with 22.5 mT/m and 200 T/m/s gradient system limits. During recordings, regular helium pump operation was maintained, while bore ventilation was turned off, resulting in ≈ 70 dB background noise.

### Signal processing

All experiments were controlled using a custom-built Labview application (National Instruments, Austin, TX, US) which contained AFG drivers and Matlab (Mathworks, Natwick, MA, US) plug-in scripts. This application was used for receiving recorded signals, processing the data, and sending control commands for the output. To diminish background noise, all recorded signals were band-pass filtered to the relevant sequence noise using a 0.3-4 kHz passband. To facilitate retrospective recording alignment, 1 s TSG blip noise was used.

### Linear time-invariant model

To predict gradient noise, a linear time-invariant (LTI) model was used. The model convolves the derivative of the gradient input *g*_*X/Y/Z*_ (*t*) with corresponding transfer functions *h*_*X/Y/Z*_ (*t*), to derive noise prediction components *p*_*X/Y/Z*_ (*t*). These are superimposed to estimate total gradient noise: 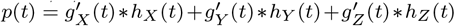. The homogeneity and superposition assumptions of the linear model were experimentally tested by pulse amplitude scaling and by comparing the superposition of single gradient pulse noise to double gradient pulses. The test results are detailed in the Supplementary Materials and Figure S1.

### Calibration sequence

A calibration pulse sequence was designed to obtain transfer functions and perform experimental error corrections (see Supplementary Materials). The sequence comprised 20 consecutive TSG pulses (triangular blips with 20 mT/m amplitude and 0.14 ms rise time, played on all coil) with TR = 3 s, interleaved with the calibration pulses with a 2 s delay. Triangular calibration gradients were played across different axes to obtain individual coil transfer functions (see Fig. S3A). The chosen gradient parameters were 20 mT/m amplitude and 0.14 ms rise time, resulting in well-characterized sound predictions in the 0.3-4 kHz range (Fig. S3B). The calibration gradients were averaged over 5 repetitions. This number was chosen based on experiments indicating plateauing simulated noise reduction under ideal playback conditions (Fig. S3C).

### Feed-forward signal corrections and evaluation metrics

For the equalizer implementation, the inverse 10001 point filter was fitted by deconvolution using a Toeplitz matrix, with 500 point non-causal element. The EQ inputs/outputs were defined as the playback and original recording of a gradient blip. To evaluate the reduction with different EQ orders, live reduction values for each EQ iteration were estimated over five repetitions for each coil gradient blip (20 mT/m amplitude and 0.14 ms rise time), over a 100 ms window.

For latency correction, the signals were over-sampled by a factor of 100, and maximum cross-correlation was used to identify the delay time. The final signal was obtained by down-sampling to the original sample rate. To evaluate the latency effects in -80 to 80 *μ*s range, 36 equally spaced steps were used. Five repetitions were used, measuring the reduction over a 100 ms window with the highest SPL.

The recorder clock-mismatch was estimated from the accumulating displacement of TSG noise clips in the calibration sequence. To evaluate the clock-mismatch correction, five live reduction repetitions of X gradient coil blips were acquired, and noise reduction was evaluated over a 100 ms window.

### Noise reduction metrics

A variety of 2D steady-state free precession (FISP) sequences were used with different sequence settings. The number of serial averages was chosen to achieve a total duration of ≈10 s for each sequence. Complete sequence parameters are listed in Supplementary Table S1.

Step 1-3 data were derived from the same 6 scanning sessions, while Step 4 data (simulated reduction under ideal timing, playback and model conditions) were acquired in a separate scanning session, where each sequence noise was acquired with 5 repetitions. For Steps 3 and 4, only one repetition of scanner-only and prediction-only noise was acquired per sequence per session. Hence, the data represents a mean over 10 sequences and 6 scanning sessions. All noise reduction values in Fig. 3 were estimated over a time window corresponding to active gradient coils (8-10 s). The frequency analysis was performed in 100 Hz bins. Pairwise p-values for low, mid-range and high frequencies (300-700, 700-1700 and 1700-4000 Hz) were derived from t-tests of band-passed live reduction (Step 1) data, averaged over all sequences and scanning sessions.

For studying robustness to sequence modifications, 2D balanced steady-state free precession (bSSFP) sequence variations were used (sequence parameters in Supplementary Table S1). Four acoustically significant sequence parameters were individually changed in four scanning sessions. Reduction values were estimated over a time window corresponding to active gradient coils (9-39 s).

For temporal stability analysis, the noise reduction cascade for the single-TR sequence was estimated by acquiring 15 noise repetitions for each step. The data is represented as the mean reduction over a 200 ms most SPL-intense window with error bars indicating ±SD in Fig. 4F. Single-TR sequence reduction as a mean over a 10 ms sliding window is represented as a mean of 15 time-aligned repetitions, with ±SD error bars.

## Data Availability

All experimental acoustic noise data, animated abstract and other supplementary videos can be found at https://gitlab.tudelft.nl/mars-lab/predictive-noise-canceling.

## Acknowledgments

The authors would like to thank Eric Verschuur for seminal discussions on acoustic wave interactions, and Henry den Bok for assistance in building the noise canceling setup.

