## Supplementary Materials for "Lowering The Acoustic Noise Burden in MRI with Predictive Noise Canceling"

### Homogeneity and superposition testing in LTI system

To verify the linearity and time-invariance assumptions in the system, homogeneity and superposition properties were experimentally verified. To assess homogeneity, triangular gradient pulses were played with 1 to 22 mT/m amplitudes at three rise times (0.14 ms, 0.34 ms and 0.54 ms). The root-mean-square (RMS) sound pressure was measured over a 100 ms window containing the majority of the noise output. Five repetitions were acquired for X, Y and Z gradient coil pulses, as well as for a simultaneous triple coil gradient pulse. In all cases, excellent linearity was confirmed (Pearson-correlation coefficient $R^2 > 0.999$, Fig. S1A).

To evaluate superposition, triangular gradient pulses (0.14 ms rise time) were played simultaneously on two gradient coils at a time. The amplitude was varied linearly between the two gradient coils in eleven steps, starting from 0 mT/m to 20 mT/m amplitude on one coil and 20 mT/m to 0 mT/m on the other coil (Fig. S1B). The double gradient noise was recorded with five averages for all three combinations of gradient coils (XY, YZ, ZX). This noise was then compared to retrospectively superimposed single gradient coil pulse noise, for each amplitude mixing ratio. Single gradient coil pulses were also acquired with five averages. Simulated reduction under ideal timing and playback conditions for the double-coil gradient pulses was then evaluated in two modes: using the superimposed single-coil gradients; and using the double-gradient noise as a baseline. For the latter, simulated reduction values excluded the pairs of identical signals. Up to 2.4 dB higher error is observed at high mixing ratios, indicating moderate superposition-induced deviations.

### Calibration and error correction integration

During the noise canceling experiments, transfer function calibration sequence (Fig. S3A) recordings were repurposed to derive the feed-forward corrections for efficiency.

Recorded TSG train noise was used to estimate the recorder-induced clock mismatch, and the signals were resampled accordingly. Using the first five TSG repetitions, an initial guess was derived for the inverse frequency distortion filter $h_{sys}^{-1}$ ($1^{st}$ order EQ filter). The recorded TSG noise was passed as an EQ input, and the output was recorded during the first five TRs of the calibration sequence. An inverse distortion filter (≈ 230 ms) was calculated in time domain using deconvolution by Toeplitz matrix construction. The subsequent noise prediction signals were convolved with $h_{sys}^{-1}$ prior to output.

After the first calibration sequence iteration, the sequence was repeated for output tuning (calibration step 2). On the scanner, only TSG pulses were played, and the calibration pulses were replaced by the noise prediction output, convolved with an inverse distortion filter. After performing the two-step recorder clock-mismatch correction, output latency was estimated by averaging the latency of five X/Y/Z pulses, relative to the scanner noise in the first calibration step.

During this step, a higher-order EQ filter was derived by using X/Y/Z noise prediction as EQ input and the corresponding recorded noise as EQ output (averaged over 5 repetitions each). $2^{nd}$ order inverse distortion filter was estimated for the three coils separately, and subsequently averaged. The total inverse frequency distortion filter $h_{sys}^{-1}$ was updated by convolving the $1^{st}$ and $2^{nd}$ order inverse distortion filters. This procedure was repeated in another iteration of calibration step 2, to achieve a $3^{rd}$ order EQ correction.

1 Department of Imaging Physics, Delft University of Technology, Lorentzweg 1, Delft, 2628 CJ, The Netherlands

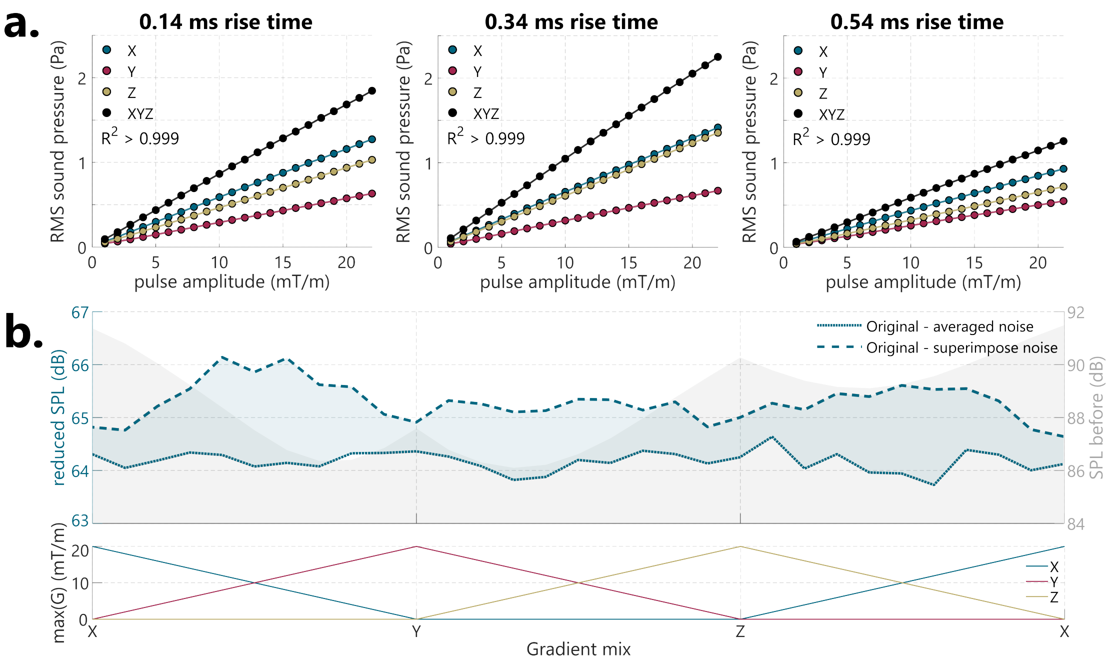


**FIGURE S1. System linearity evaluation. a**, linearity test - total root mean square (RMS) sound pressure measured for triangular gradients for each gradient coil separately or all three gradient coils combined. The gradient amplitude was varied for 0.14/0.34/0.54 ms rise times. Excellent linearity is observed for each gradient coil and rise-time ($R^2 > 0.999$). **b**, superposition test. For linear combinations of two-coil triangular gradients (0.14 ms rise time), the reduction modes were simulated under ideal timing and playback conditions, and using two models: retrospectively superimposed averaged single-coil gradient noise (dashed line) and averaged double-coil gradient noise (dotted line, equivalent to Step 4). The blue shaded area represents the superposition error that reaches up to 2.4 dB at high mixing ratios, indicating moderate deviations from the linear time invariant (LTI) model. The gray shaded area represents gradient SPL before simulated noise reduction.

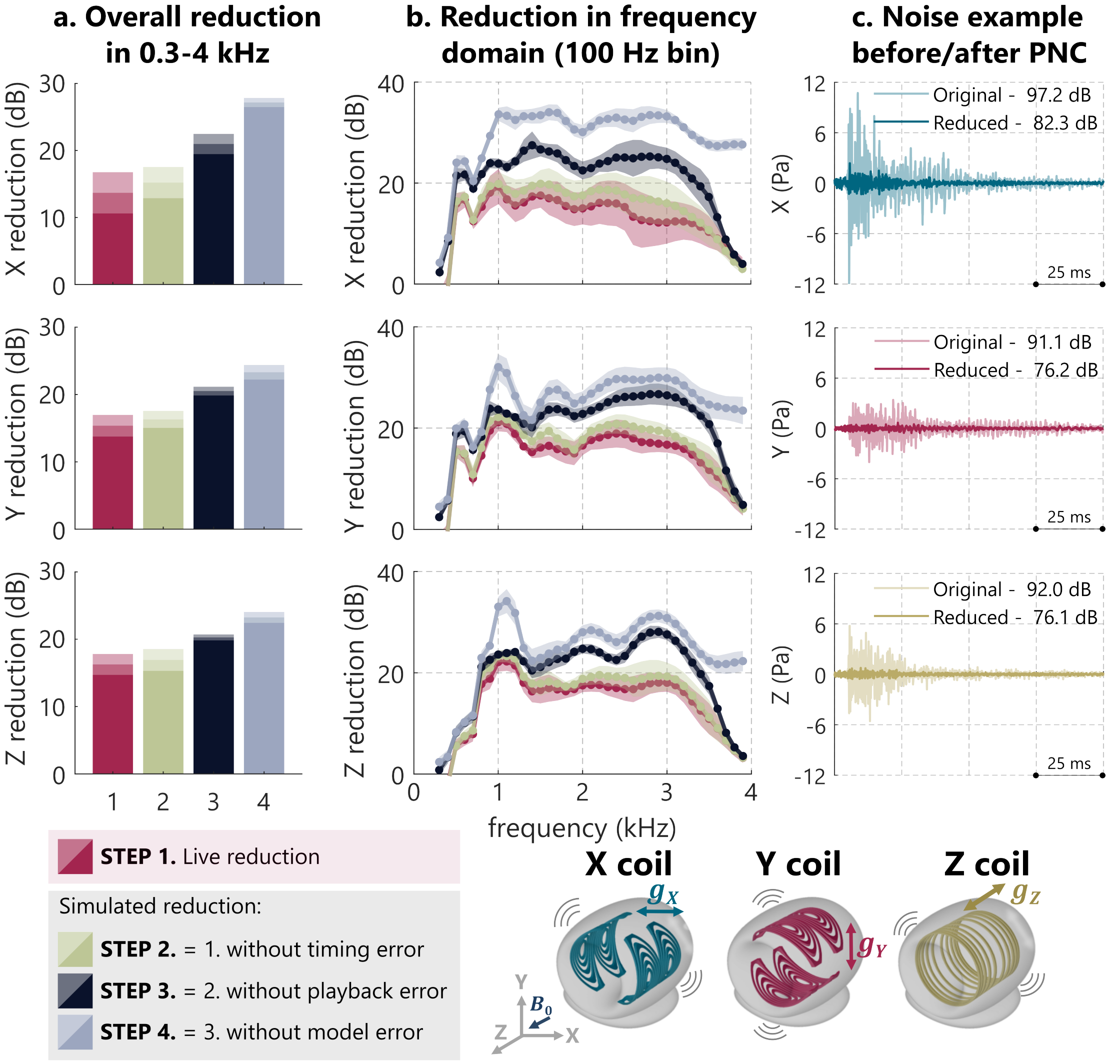


**FIGURE S2. Triangular pulse reduction cascade for X/Y/Z gradient coils. a**, triangular gradient pulse live noise reduction and simulated reduction when virtually excluding experimental error sources in the four steps for the wider frequency range of 0.3-4 kHz. ± standard deviation is indicated as shading in the error bars. **b**, Step 1-4 reduction cascade in the frequency domain over 100 Hz bins. **c**, example live noise reduction (Step 1) in the time domain for a single triangular gradient pulses.

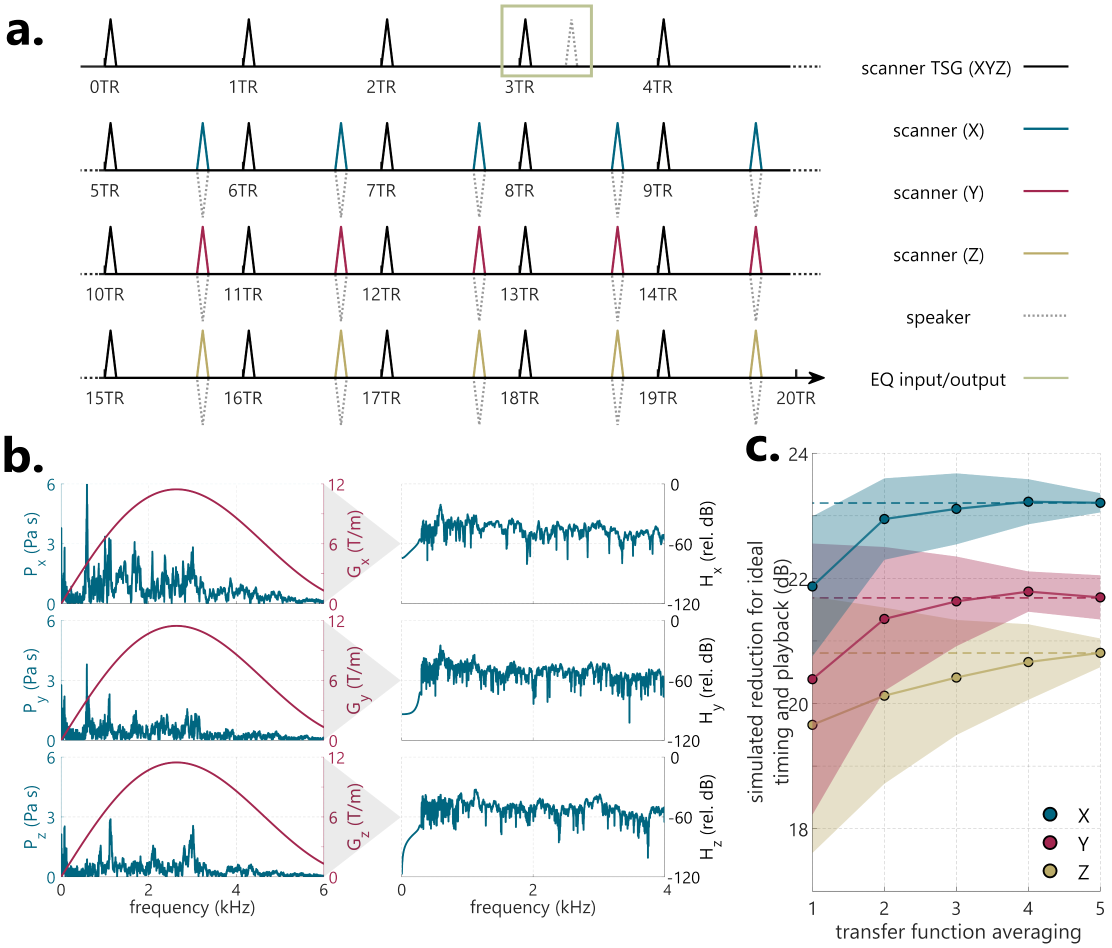


**FIGURE S3. Calibration sequence scheme details.** **a**, pulse sequence diagram. Each repetition time (TR = 3 s) begins with TTL synchronized gradient (TSG, black line). The first 5 start-up TRs are used for EQ calibration. In all subsequent TRs, the TSG is followed by a gradient pulse on a single gradient coil (X/Y/Z) after 2 s delay. **b**, exemplary noise spectrum and gradient input spectrum, used to derive the acoustic transfer functions (right) in the frequency domain. **c**, simulated noise reduction under ideal timing and playback conditions (Step 3) for X/Y/Z pulses with an increasing number of averages. The dotted line represents the plateau-approaching line at five averages.

**TABLE S1. MRI sequence parameter list.** FISP - fast imaging with steady-state free precession; bSSFP - balanced steady state free precession; EPI - echo-planar imaging; TR - repetition time; TE - echo time; BW - bandwidth; FOV - field of view; AP/RL/FH - angulation values from orientation plane - anterior-posterior/right-left/feet-head.

| **Experiment name** | **sequence type** | **TR (ms)** | **TE (ms)** | **BW (Hz/px)** | **FOV ($mm^3$)** | **voxel ($mm^3$)** | **plane** | **ang.(°AP/ RL/FH)** |
|---|---|---|---|---|---|---|---|---|
| **Sequence 1** | 2D FISP | 15 | 8 | 913.2 | 230×230×1 | 1×1×1 | coronal | 0/0/0 |
| **Sequence 2** | 2D FISP | 7.6 | 3.7 | 189.7 | 230×230×10 | 1×1×10 | transverse | 14/-27/12 |
| **Sequence 3** | 2D FISP | 9.7 | 5.7 | 918.3 | 330×330×1 | 1×1×1 | transverse | 30/25/10 |
| **Sequence 4** | 2D FISP (22 shots) | 5.1 | 3.4 | 826.2 | 220×220×5 | 0.9×0.9×5 | saggital | 0/0/0 |
| **Sequence 5** | 2D FISP (80 shots) | 18 | 3 | 473.5 | 200×200×10 | 0.5×0.5×10 | saggital | 30/25/10 |
| **Sequence 6** | 2D FISP | 25 | 11 | 86.9 | 212×212×1 | 2×2×1 | coronal | 5/29/10 |
| **Sequence 7** | 2D FISP with flow comp.; EPI factor 3 | 20 | 3.7 | 913.2 | 230×230×20 | 1×1×20 | saggital | 45/2/34 |
| **Sequence 8** | 2D FISP | 14 | 1.07 | 2790.2 | 330×330×5 | 3×3×5 | coronal | 30/25/17 |
| **Sequence 9** | 2D FISP (22 shots) | 27 | 1.07 | 2790.2 | 330×330×5 | 3×3×5 | coronal | 30/25/10 |
| **Sequence 10** | 2D FISP with flow comp.; EPI factor 3 | 44 | 29 | 35 | 230×230×10 | 1×1×10 | coronal | 0/0/0 |
| **Slice thickness study** | 2D bSSFP | 12 | 5.7 | 757.6 | 400×400× **(0.5,1:10)** | 0.8×0.8× **(0.5,1:10)** | coronal | 30/36/-20 |
| **Bandwidth study** | 2D bSSFP | 9.3 | 4.6 | **A*** | 400×400×10 | 0.8×0.8×10 | coronal | 30/36/-20 |
| **Rep. time study** | 2D bSSFP | **B*** | 2.1 | 757.6 | 400×400×10 | 0.8×0.8×10 | coronal | 30/36/-20 |
| **Rotation study** | 2D bSSFP | 12 | 5.7 | 217 | 400×400×10 | 1×1×10 | coronal | 10/-15/ **-142:18:20** |
| **Long seq. 1, single-TR** | 2D bSSFP | 250 | 5.7 | 757.6 | 400×400×10 | 0.8×0.8×10 | transverse | 0/0/0 |
| **Long seq. 2** | 2D bSSFP | 12 | 5.7 | 757.6 | 400×400×10 | 0.8×0.8×10 | transverse | 0/0/0 |
| **Long seq. 3** | 3D bSSFP; 22 slices | 12 | 5.7 | 757.6 | 400×400×1 | 0.8×0.8×1 | transverse | 0/0/0 |

***A** - 271.7, 308.6, 347.2, 387.6, 427.4, 463.0, 505.1, 543.5, 581.4, 617.3 Hz/px
***B** - 6.5, 10.8, 17.2, 34.4, 68.8, 137.6, 275.2, 550.4 ms
